# Crowdfunding Medical Care: A Comparison of Online Medical Fundraising in Canada, the United Kingdom, and the United States

**DOI:** 10.1101/2020.03.26.20044669

**Authors:** Sameh N. Saleh, Ezimamaka Ajufo, Christoph U. Lehmann, Richard J. Medford

## Abstract

**Background:** Medical crowdfunding is increasingly used to finance personal healthcare costs in Canada (CAN), United Kingdom (UK), and United States (US) despite major differences in their healthcare systems. Yet, it lacks comparative descriptive research to guide policy changes that can promote equitable and accessible healthcare.

**Methods:** We conducted a cross-sectional analysis of Canadian, British, and American campaigns between February 2018 and March 2019 on the GoFundMe platform (n=3,396). We extracted and manually reviewed variables from campaigns on each country’s GoFundMe discovery webpage, explored campaign characteristics, and compared each country’s campaign demographics to its respective national census. We fit multivariate linear regression models for funds raised for the cohort and for each country.

**Results:** We examined 1,091 Canadian, 1,082 British, and 1,223 American campaigns. US campaigns (median [IQR] $38,204 [$31,200 to $52,123]) raised more funds than those in CAN ($12,662 [$9,377 to $19,251]) and the UK ($6,285 [$4,028 to $12,348]). Female (38.4% of campaigns vs. 50.9% of US census; p<0.001) and black (5.3% of campaigns vs. 13.4% of US census; p<0.001) beneficiaries were underrepresented in US campaigns. In the full cohort, blacks raised $4,007 less (95% confidence interval [CI] -$6,913 to -$1,101; p=0.007) and males raised $1,742 more (95% CI $583 to $2,901; p=0.003) per campaign. Cancer was the most common diagnosis represented overall (54.5%). Across all diagnoses, campaigns primarily for routine treatment expenses were three times more common in the United States compared to Canada and the United Kingdom (CAN 21.9% vs. UK 26.6% vs. US 77.9%; p<0.001). However, campaigns with routine care were less successful overall, raising $4,589 less per campaign (CI -$6,429 to - $2,749; p<0.001). Campaigns primarily for alternative treatment expenses were nearly five times as common for cancer (24%) than for non-cancer (5%) diagnoses.

**Discussion:** The trends observed suggest that there are important gaps in healthcare provision in all of the countries examined across a wide range of diagnoses. Although medical crowdfunding has the potential to provide short-term relief from medical financial burden for a privileged subset of patients, it may carry wider-reaching adverse societal effects including the promotion of racial and gender disparities in healthcare. Further work is needed to inform policy changes that promote equitable and accessible healthcare through this practice.

**Funding:** None.

## Introduction

Crowdfunding, the online solicitation of public donations, has become an important form of financing to pay for accumulated personal healthcare debts. Approximately a third of all crowdfunding campaigns are intended to pay for healthcare-related costs.^1–3^ The growing importance of medical crowdfunding (MCF) is reflected by trends on GoFundMe, the largest social crowdfunding platform in the world.^3,4^. In 2011, medical causes raised $1.6 million GoFundMe; in 2014, the amount had increased almost a hundredfold to $150 million and in 2016, more than $650 million.^1,3^

The growing reliance of United States (US) healthcare consumers on MCF is primarily attributed to increasing healthcare costs and the lack of a publicly funded healthcare system.^1,5,6^ However, the popularity of MCF in developed countries with universal healthcare such as Canada (CAN) and the United Kingdom (UK) suggests additional reasons.^7–10^ MCF has financed an array of experimental and alternative therapies as well as gaps in public services.^11–14^ Despite its growth, there is growing, but limited empirical research on MCF, including research on socio-demographics of beneficiaries and the diagnoses and treatments championed. Moreover, inequity, barriers to access, invasion of privacy, fraud, and dangerous therapies have all been associated with MCF, but are poorly understood.^4,7,15–19^

The goal of this study was to evaluate three important aspects of MCF in different healthcare systems: the cause for turning to crowdfunding, characteristics of beneficiaries and campaigns, and factors associated with funding success. We selected GoFundMe as an ideal environment to study. As of 2018, the platform reportedly controlled 90% of the social crowdfunding market in the US and 80% of the global market.^20^ We studied consecutive campaigns from GoFundMe in CAN, the UK, and the US, the three countries with the largest markets on this platform.

## Methods

### Study Population

We conducted a cross-sectional analysis of campaigns launched between February 2018 and March 2019 from the GoFundMe domains in CAN, the UK, and the US. GoFundMe offers 21 cause categories for users to select; using a web scraping tool (Beautiful Soup^21^), we extracted campaigns from the “Medical” category only. For each country’s GoFundMe domain, we accessed the 1,000 available medical campaigns on the “Discover” page (www.gofundme.com/discover/medical-fundraiser) at two timepoints in February 2019 and again 30 days later in March 2019. Our query resulted in 1,107, 1,117, and 1,232 unique campaigns in CAN, the UK, and the US, respectively (Figure 1). We excluded 61 campaigns that did not benefit a unique patient such as those that raised funds for a general cause, research, or non-profit organization.

**Figure 1.**
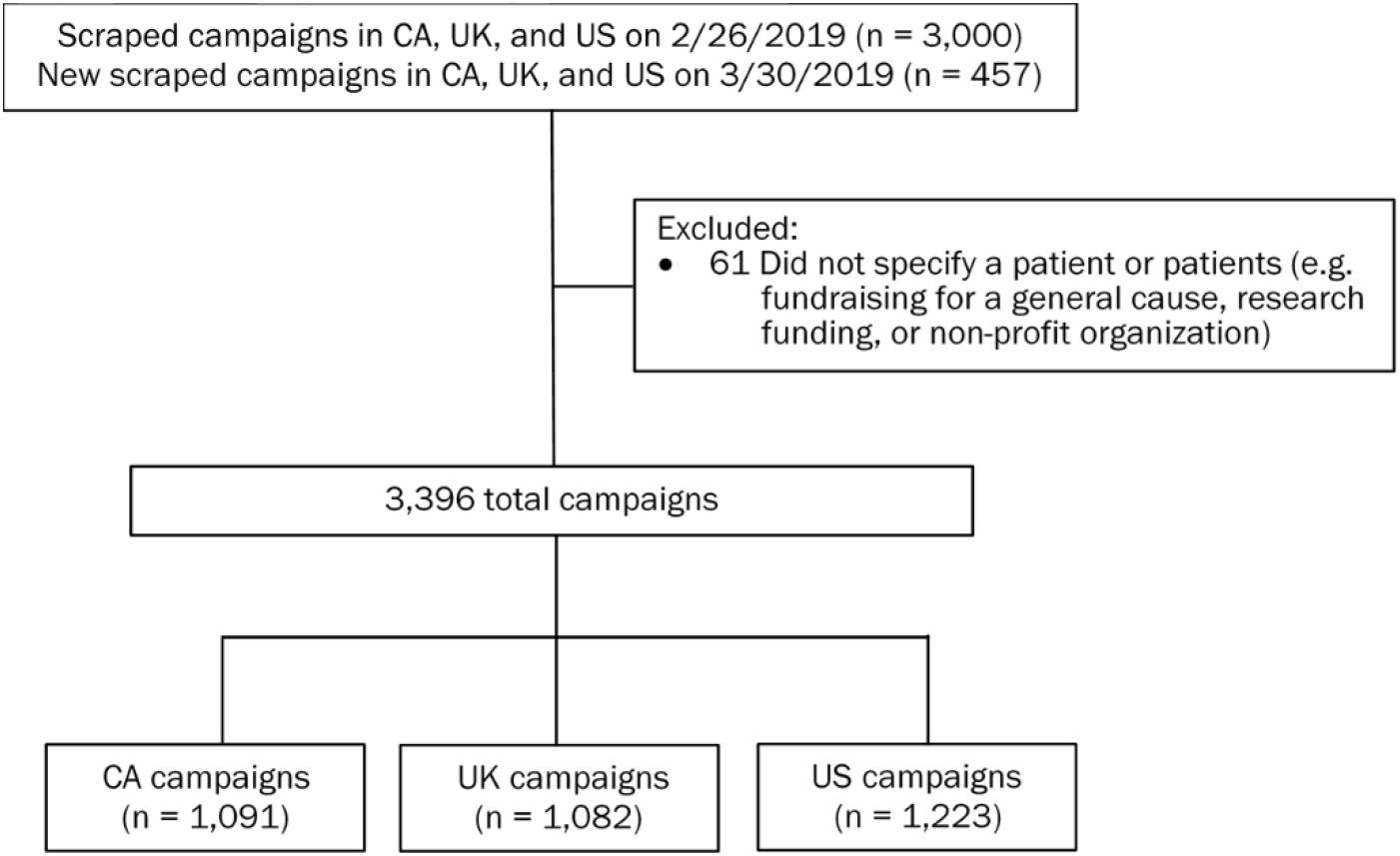
Study flow diagram.

### Study Variables

For each campaign, we extracted the quantitative data displayed on the campaign webpage including the monetary goal, amount raised to date, number of donors, location, length of the fundraising campaign, Facebook shares, and GoFundMe hearts (the equivalent of webpage “likes”). Data were complete except Facebook shares, with 2.6% of campaigns missing values, for which we imputed a value of zero. We converted all funds to US dollars based on currency exchange rates on the day we accessed the data.

Using the text and media on each initial campaign post, a two-person manual review of the campaigns collected data on demographics, diagnosis, type of treatment (routine, experimental [i.e., not yet approved], approved but inaccessible [i.e., unavailable in the patient’s location], alternative [i.e., treatments used in lieu of standard care], and unspecified), funding intent of the campaign (primarily for treatment costs), patient location (residing outside of the campaign country), and status (alive or deceased). eAppendix in the Supplement details the variables and definitions used for labeling the columns. Each individual reviewed 55% of the data with 10% overlap among the reviewers. Concordance analysis showed the inter-rater reliability (κ) exceeded 0.77 for all but one category (eTable 1 in the Supplement). A κ greater than 0.8 indicates almost perfect agreement.^22^ There was greater than 89.2% raw agreement in all categories, with most greater than 98%. A blinded third reviewer adjudicated any discrepancies between the two reviewers.

**Table 1.**
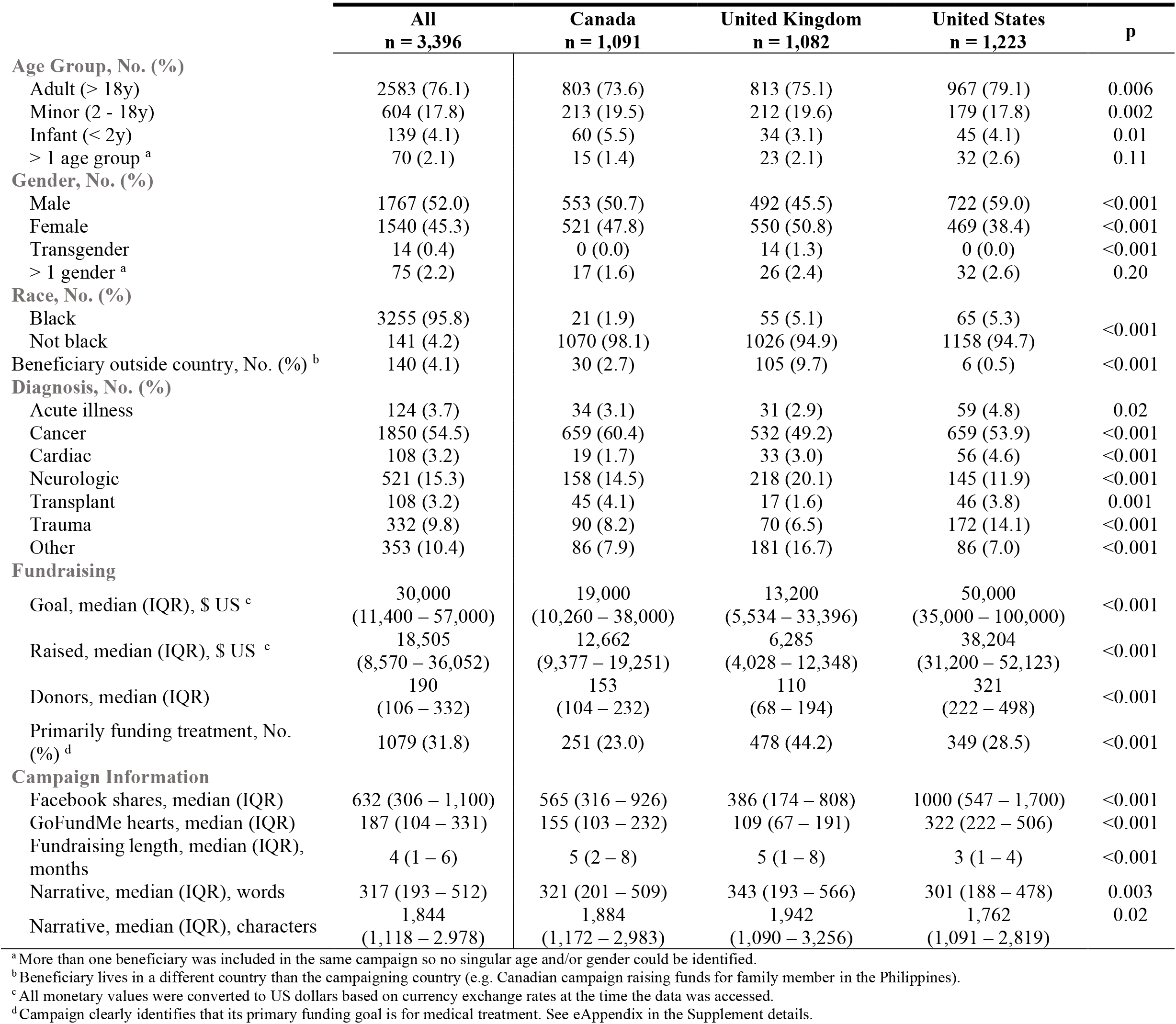
Baseline characteristics stratified by country for entire cohort. Continuous variables presented as median (IQR), categorical variables as number (%).

### Statistical Analysis

SS completed all analysis using Python software, version 3.7.2 (Python Software Foundation). We performed descriptive data analysis of all variables to evaluate trends and common characteristics of MCF in the three countries. Given the non-Gaussian populations, we used Kruskal-Wallis, Chi-squared, and Fischer’s exact testing to detect statistical differences among the three groups. To test for representativeness, we compared the campaign demographics to respective national census data (2016 CAN census^23^, 2018 UK estimate^24^, 2018 US estimate^25^) using Chi-squared testing.

We performed multivariable linear regressions for the full cohort and by country using funds raised as the prespecified primary outcome and completed a sensitivity analysis for other outcomes. Funds raised has been a common outcome reported and used in MCF research.^11,12^ We constructed a pairwise Pearson correlation matrix among all variables and excluded repetitive variables with high collinearity from the analysis resulting in the removal of GFM hearts (“likes”) and campaign narrative character count from the analysis due to high correlation with the number of donors and narrative word count, respectively. To avoid collinearity in categorical variables, we dropped the least specific variable (e.g. “Other” for diagnosis or “Unspecified” for the treatment type). We further removed the most collinear variables (e.g. “Female” given high collinearity with “Male”). The list of variables included is displayed in Table 3. We evaluated the models using the R^2^ coefficient and compared variable coefficients with confidence intervals to determine the strength of effect and statistical significance. We did not adjust for multiple comparisons as this was an exploratory study and should be interpreted as hypothesis-generating. Inferences may not be reproducible and further dedicated studies are needed to confirm the results.

**Table 2.** Comparison of the proportion of campaign representation to the proportion of population according to national census data (2016 CAN, 2018 UK, 2018 US). Campaigns with more than one beneficiary were excluded.

|  | Canada |  |  | United Kingdom |  |  | United States |  |  |
| --- | --- | --- | --- | --- | --- | --- | --- | --- | --- |
|  | Campaign<br>n (%) | Census<br>% | p<sup>a</sup> | Campaign<br>n (%) | Census<br>% | p<sup>a</sup> | Campaign<br>n (%) | Census<br>% | p<sup>a</sup> |
| **Age Group, No. (%)** |  |  |  |  |  |  |  |  |  |
| Adult (> 18y) | 803 (74.8) | 80.0 | <0.001 | 813 (76.8) | 78.8 | 0.11 | 967 (81.2) | 77.6 | 0.003 |
| Minor (2 - 18y) | 213 (19.8) | 17.9 | 0.11 | 212 (20.0) | 18.9 | 0.36 | 179 (15.0) | 20.1 | <0.001 |
| Infant (< 2y) | 60 (5.6) | 2.1 | <0.001 | 34 (3.2) | 2.3 | 0.03 | 45 (3.8) | 2.4 | 0.002 |
| **Gender, No. (%)** |  |  |  |  |  |  |  |  |  |
| Male | 553 (51.5) | 49.1 | 0.13 | 492 (47.2) | 49.3 | 0.18 | 722 (60.6) | 49.2 | <0.001 |
| Female | 521 (48.5) | 50.9 |  | 550 (52.8) | 50.7 |  | 469 (39.4) | 50.8 |  |
| **Race, No (%)** |  |  |  |  |  |  |  |  |  |
| Black | 21 (1.9) | 3.5 | 0.007 | 55 (5.1) | 3.1 | <0.001 | 65 (5.3) | 13.4 | <0.001 |
| Not black | 1070 (98.1) | 96.5 |  | 1026 (94.9) | 96.9 |  | 1158 (94.7) | 86.6 |  |

<sup>a</sup>The p-value is determined by using Chi-squared testing to compare the proportion of campaign representation to the proportion of the national census in each country.

**Table 3.** Multivariable linear regression for amount raised for the full cohort and for each country. Values reflect linear regression coefficient and 95% CI for each variable after adjustment for all other variables listed in the table.

|  |  | Linear Regression Coefficients (95% CI) |  |  |  |  |  |  |  |
| --- | --- | --- | --- | --- | --- | --- | --- | --- | --- |
|  |  | All<br>(R<sup>2</sup> = 0.67) | p | Canada<br>(R<sup>2</sup> = 0.40) | p | United Kingdom<br>(R<sup>2</sup> = 0.68) | p | United States<br>(R<sup>2</sup> = 0.53) | p |
| Demographics/Location |  |  |  |  |  |  |  |  |  |
|  | Male | 1,742<br>(583 to 2,901) | 0.003 | 2,914<br>(1,277 to 4,552) | <0.001 | 384<br>(-1,699 to 2,680) | 0.72 | 1,733<br>(-412 to 3,878) | 0.11 |
|  | Transgender | -2,129<br>(-11,200 to 6, 902) | 0.65 | N/A | -- | -4,031<br>(13,300 to 5,267) | 0.40 | N/A | -- |
|  | Adult | 780<br>(-674 to 2,234) | 0.29 | 193<br>(-1,867 to 2,254) | 0.85 | 28.4<br>(-2,456 to 2,513) | 0.98 | 1,238<br>(-1,623 to 4,098) | 0.40 |
|  | Infant | -1,800<br>(-4,908 to 1,307) | 0.26 | -2,857<br>(-6,813 to 1,099) | 0.16 | -3,027<br>(-9,142 to 3,088) | 0.33 | 2,053<br>(-4,012 to 8,118) | 0.51 |
|  | Black | -4,007<br>(-6,913 to -1,101) | 0.007 | -4,639<br>(-10,500 to 1,232) | 0.12 | -4,454<br>(-9309 to 400) | 0.07 | -2,879<br>(-7,606 to 1,849) | 0.23 |
|  | Canadian campaign | -2,419<br>(-3,757 to -1,081) | <0.001 | N/A | -- | N/A | -- | N/A | -- |
|  | UK campaign | -6,769<br>(-8,142 to -5,396) | <0.001 | N/A | -- | N/A | -- | N/A | -- |
|  | US campaign | 16,930<br>(15,500 to 18,400) | <0.001 | N/A | -- | N/A | -- | N/A | -- |
|  | Non-Medicaid expansion | N/A | -- | N/A | -- | N/A | -- | 1,469<br>(-933 to 3,870) | 0.23 |
|  | Beneficiary outside country <sup>a</sup> | -70.9<br>(-3,185 to 3,043) | 0.96 | -3,042<br>(-8284 to 2,200) | 0.26 | 676<br>(-3,476 to 4,829) | 0.75 | -5,853<br>(-20,900 to 9,154) | 0.44 |
|  | Beneficiary deceased | -3,031<br>(-6,889 to 828) | 0.12 | 2,315<br>(-4,345 to 8,975) | 0.50 | -2,620<br>(-9,635 to 4,396) | 0.46 | -6,121<br>(-12,300 to 69.2) | 0.05 |
| Diagnosis |  |  |  |  |  |  |  |  |  |
|  | Acute illness | -1,697<br>(-5,719 to 2,325) | 0.41 | -2,579<br>(-7,650 to 2,492) | 0.32 | -433<br>(-6,389 to 5,523) | 0.89 | -4,471<br>(-10,600 to 1,658) | 0.15 |
|  | Cancer | 2,379<br>(-1,121 to 5,878) | 0.18 | 1,593<br>(-1,347 to 4,533) | 0.29 | 340<br>(-2,556 to 3,236) | 0.82 | 1,726<br>(-3,474 to 6,925) | 0.52 |
|  | Cardiac | 3,799<br>(-215 to 7,812) | 0.06 | 806<br>(-5,624 to 7,236) | 0.81 | 2,349<br>(-3,888 to 8,586) | 0.46 | 4,517<br>(-1,551 to 10,600) | 0.14 |
|  | Neurologic | 2,389<br>(-1,106 to 5.884) | 0.18 | -1,000<br>(-4,319 to 2,319) | 0.56 | 1,613<br>(-1,666 to 4,892) | 0.34 | 4,350<br>(-1,063 to 9,762) | 0.12 |
|  | Transplant | -451<br>(-4,214 to 3,312) | 0.81 | -1,704<br>(-6,114 to 2,705) | 0.45 | 1,885<br>(-5,650 to 9,421) | 0.62 | 316<br>(-5,088 to 5,720) | 0.91 |
|  | Trauma | 1,156<br>(-2,720 to 5,033) | 0.56 | 990<br>(-3,048 to 5,027) | 0.63 | -915<br>(-5,587 to 3,756) | 0.70 | 2,343<br>(-3,343 to 8,029) | 0.42 |
| Treatment Type/Details |  |  |  |  |  |  |  |  |  |
|  | Alternative | -1,709<br>(-4,156 to 739) | 0.17 | -979<br>(-4,448 to 2,489) | 0.58 | -126<br>(-4,330 to 4,078) | 0.95 | -5,384<br>(-10,200 to -525) | 0.03 |
|  | Approved – not accessible | 23<br>(-2,852 to 2,899) | 0.99 | 3,247<br>(-953 to 7,447) | 0.13 | -1,238<br>(-5,315 to 2,840) | 0.55 | 17,390<br>(-1,185 to 36,000) | 0.07 |

| Linear Regression Coefficients (95% CI) |  |  |  |  |  |  |  |  |
| --- | --- | --- | --- | --- | --- | --- | --- | --- |
|  | All<br>(R<sup>2</sup> = 0.67) | p | Canada<br>(R<sup>2</sup> = 0.40) | p | United Kingdom<br>(R<sup>2</sup> = 0.68) | p | United States<br>(R<sup>2</sup> = 0.53) | p |
| Experimental – not approved | 2,651<br>(90 to 5,213) | 0.04 | 1,609<br>(-2,034 to 5,252) | 0.39 | 4,631<br>(161 to 9,101) | 0.04 | -487<br>(-5,614 to 4,640) | 0.85 |
| Routine care | -4,589<br>(-6,429 to -2,749) | <0.001 | -1,994<br>(-4,562 to 574) | 0.13 | -1,323<br>(-4,471 to 1,824) | 0.41 | -11,060<br>(-14,800 to -7,358) | <0.001 |
| Experimental stem cell <sup>b</sup> | -200<br>(-4,680 to 4,280) | 0.93 | -713.8<br>(-7,332 to 5,904) | 0.83 | -2,731<br>(-9,026 to 3,564) | 0.40 | 14,310<br>(1,026 to 27,600) | 0.04 |
| **Campaign Information & Social Media** |  |  |  |  |  |  |  |  |
| Goal, \$ US <sup>c</sup> | 0.037<br>(0.031 to 0.042) | <0.001 | 0.092<br>(0.074 to 0.110) | <0.001 | 0.041<br>(0.027 to 0.054) | <0.001 | 0.029<br>(0.022 to 0.036) | <0.001 |
| Fundraising length, months | 329<br>(141 to 518) | 0.001 | 152.0<br>(-81.2 to 385.2) | 0.20 | 490<br>(208 to 771) | 0.001 | 267<br>(-336 to 871) | 0.39 |
| Primarily funding treatment <sup>d</sup> | 937<br>(-597 to 2,470) | 0.23 | 1,522<br>(-1,116 to 4,159) | 0.26 | 2,308<br>(-725 to 5,340) | 0.14 | -234<br>(-2,661 to 2,193) | 0.85 |
| Narrative, words | 1.626<br>(0.002 to 3.249) | 0.05 | 0.703<br>(-1.479 to 2.885) | 0.53 | 0.709<br>(-2.247 to 3.665) | 0.64 | 3.777<br>(0.686 to 6.867) | 0.02 |
| Donors, n | 43.28<br>(41.33 to 45.24) | <0.001 | 32.04<br>(27.73 to 36.35) | <0.001 | 50.59<br>(46.47 to 54.70) | <0.001 | 39.86<br>(36.98 to 42.74) | <0.001 |
| Facebook shares | -0.346<br>(-0.925 to 0.234) | 0.24 | 1.026<br>(-0.543 to 2.596) | 0.20 | 0.005<br>(-1.462 to 1.473) | 0.99 | -0.744<br>(-1.499 to 0.010) | 0.05 |

<sup>a</sup> Beneficiary of the campaign lives in a different country than a campaigning country (e.g Canadian campaign raising funds for family member in the Philippines).
<sup>b</sup> Campaign mentions stem cell treatment or transplant in an experimental application (i.e. not for leukemias, lymphomas, etc.).
<sup>c</sup> All monetary values were converted to US dollars based on currency exchange rates at the time the data was accessed.
<sup>d</sup> Campaign clearly identifies that its primary funding goal is for medical treatment. See eAppendix in the Supplement for details.

## Results

### Beneficiary Demographics and Diagnoses

Of the 3,396 campaigns, 1,091 originated in CAN, 1,082 in the UK, and 1,223 in the US. Table 1 presents the campaign characteristics, stratified by country. Table 2 compares the campaign demographics in each country to its national census. Most campaign beneficiaries were male (52.0%), adult (76.1%), and non-black (95.8%). The US had the highest proportion (59.0%) of male beneficiaries (CAN vs. US and UK vs. US, both p<0.001). Females comprise 50.9% of the US population, but were beneficiaries in only 38.4% of US campaigns (p<0.001). In the US, adults were over-represented compared to the census (p=0.003). The US had more black beneficiaries than CAN and a similar proportion to the UK. However, compared to national census representations, black beneficiaries were most underrepresented in the US (5.3% vs. 13.4%, p<0.001). Black Canadians were also underrepresented (1.9% vs. 3.5%, p=0.004), while blacks in the UK were overrepresented in campaigns (5.1% vs. 3.3%, p<0.001). Cancer was the most common diagnosis represented in fundraisers (54.5%), followed by neurologic (15.3%), other (10.4%), and trauma (9.8%). Compared to CAN and the UK, the US proportionally had the most campaigns for acute illness, cardiac, and trauma.

### Fundraising Characteristics

Campaigns in this study collectively raised $92.9 million accounting for 51.9% of the total funds sought. Only 33.3% of all campaigns had met their goal at data extraction. Funds raised ranged from $2,772 to $343,762 per campaign. US campaigns set higher fundraising goals than those in CAN and the UK. Accounting for $57.8 million (62.2% of the total funds raised), US campaigns raised approximately three times the funds of CAN campaigns (p<0.001) and six times the funds of UK campaigns (p<0.001) on average, despite being two months shorter in duration (p<0.001). US campaigns had significantly more donors and Facebook shares (all p<0.001).

### Treatment Type

Figure 2 summarizes the treatment types for which campaigns raised funding, stratified by country. While routine care was the most common treatment type, accounting for 69.4% of campaigns, 84% of US campaigns focused on routine care compared to 69% of CAN campaigns and 54% of UK campaigns (CAN vs. US and UK vs. US, both p<0.001) (Figure 2A).

**Figure 2.**
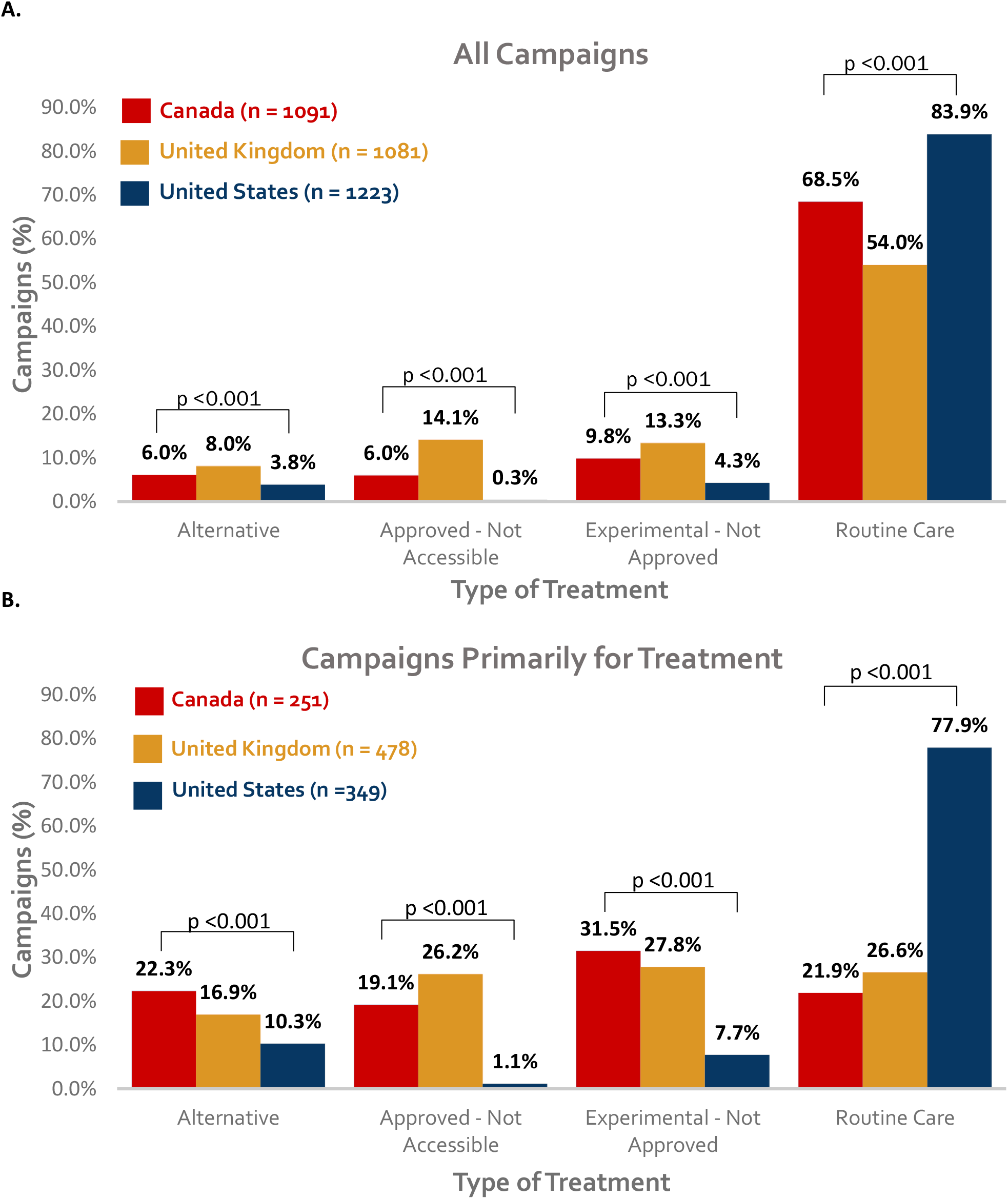
Primary type of treatment by country for A) all campaigns and B) for campaigns primarily for treatment costs.

Experimental, approved but inaccessible, and alternative care were all more common among CAN and UK campaigns compared to US campaigns. The UK had substantially more approved but inaccessible care than CAN (p<0.001) or the US (p<0.001). Approximately a third (n=1,078) of all campaigns were fundraising primarily for treatment expenses. Of these, campaigns funding routine care were approximately three times more common in the US than CAN or the UK (CAN 21.9% vs. UK 26.6% vs. US 77.9%, p<0.001) (Figure 2B). Campaigns primarily for treatment expenses in CAN and the UK were similarly distributed among routine, alternative, experimental, and unavailable therapies. Nearly a fourth of cancer campaigns primarily funding treatment were for alternative therapies, while all non-cancer diagnoses combined had less than 5% (eTable 2 in the Supplement).

### Predictors of Campaign Success

Findings from multivariable regression analysis of funds raised per campaign are presented in Table 3. Campaign country was the strongest predictor of the amount raised for the full cohort. US campaigns raised $16,930 more on average (p<0.001). Campaigning from CAN and the UK yielded $2,419 and $6,769 fewer funds, respectively (p<0.001 for both). Number of donors and fundraising goal were strongly associated with funding success. Facebook shares were not associated with funding success in the main regression model, but when donor numbers (moderately collinear with Facebook shares) were removed from the model, Facebook shares became associated with funds raised (p<0.001) [data not shown].

Beneficiary race and gender were important predictors of funds raised. Overall, black beneficiaries raised $4,007 less per campaign (p=0.007). There was a concordant but non-significant trend in the country analyses. Male beneficiaries raised $1,742 more per campaign than their counterparts (p=0.003); this trend was most pronounced in CAN where males raised $2,914 more (p<0.001). Campaigns for routine care raised $4,589 less per campaign (p<0.001); this association was strongest in the US with $11,060 less per campaign (p<0.001).

## Discussion

We examined 3,396 medical campaigns from CAN, the UK and the US, the three largest crowdfunding markets on GoFundMe, to characterize MCF beneficiaries, impetus for use of MCF, and determinants of funding success. To our knowledge, this is the largest quantitative analysis of the MCF landscape to date. US campaigns set higher goals and raised several-fold more funds than campaigns in CAN or UK. However, approximately two-thirds of campaigns in each country did not meet their funding goals. In the US, nearly 80% of campaigns primarily funding treatment were for routine care, while in CAN and the UK, funding for routine care was sought about as frequently as for alternative, approved but inaccessible, and experimental therapies. Campaigns for routine care were associated with less crowdfunding success, while campaigns for experimental therapies raised more funds. Finally, we observed gender and racial inequities among beneficiaries.

### Beneficiary demographics and inequities in crowdfunding

Crowdfunding was initially heralded as a “digital safety net” or a mechanism for democratizing charity where anyone could benefit.^7^ However, anecdotal and empirical evidence suggests that crowdfunding may exacerbate socioeconomic inequities.^9,15,26^ In our study, blacks and females were under-represented in US campaigns and blacks were under-represented in CAN campaigns. In another study of 637 randomly sampled US MCF campaigns from GoFundMe, non-white beneficiaries were also significantly under-represented, constituting only 19% of the sample while this group represents 27% of the US census.^27^ Within our sample, females and black beneficiaries raised approximately $4,000 and $1,700 less than their male and non-black counterparts, respectively. Similarly, in a US study of 850 campaigns for organ transplantation, females had raised 27% less than males after multivariable adjustment.^12^ In a CAN study of 319 campaigns, being a visible ethnic minority was associated with raising 15% less in funds, before adjustment for technological competency (using quantity of campaign images, videos, updates, and perks) and 6% less after adjustments.^7^

Race and gender disparities reflect the pillars on which MCF is dependent: access to technology, literacy, social capital, and perception. Those with socioeconomic disadvantage suffer from the “digital divide” that limits online participation due to a lack of access to information technology (e.g., computer and internet). Writing, media, and healthcare literacies, which reflect socioeconomic privilege, enable an individual to augment his/her “illness narrative”, communicate “deservingness”, and thereby, generate campaign appeal and influence^4^.

Furthermore, broader, more affluent social communities and larger social media networks are well-recognized determinants of crowdfunding success.^4,7,12^ Finally, conscious and unconscious systemic racial and gender biases likely obscure perception of worthiness in MCF campaigns.^28^ Our findings substantiate concerns that MCF facilitates the distribution of resources according to biases and preferences favoring an already privileged group of individuals and thus, contribute to widening social inequities.^29,30^

### Reasons individuals turn to medical crowdfunding and gaps in healthcare funding

If trends in MCF reflect healthcare cost coverage and insurance^7,30^, this study highlights several country-specific shortcomings in healthcare funding. The trends seen in the US suggest that its greatest systemic funding failure lies in the provision of routine health care. Despite a decrease in the uninsured rate after passage of The Affordable Care Act (ACA), as of 2018, 11.1% of adults under the age of 65 were uninsured^31^ and 29% were estimated to be underinsured^32^ (i.e., out-of-pocket costs or deductibles comprising 5-10% of their income) reflected in the over-representation of adult campaigns in the US. Most US campaigns across all diagnoses had routine care suggesting pervasive insufficient insurance coverage in the US. Diagnosis groups with more routine care such as trauma and acute illness were more common in the US. For campaigns primarily funding treatments, 78% were for routine care in the US compared to approximately 25% of analogous campaigns in both CAN and the UK. Yet, campaigns for routine care raised $11,060 less in the US, perhaps reflecting the saturation of the US MCF market with this type of campaign. US healthcare costs are the highest in the developed world and health insurance deductibles have increased eight times as much as wages since 2008.^33^ The disproportionate solicitation of MCF in the US overall and specifically for routine care likely reflect the high out-of-pocket costs associated with essential healthcare in the US.

The trends seen in CAN and UK reflect the unique failures of publicly funded healthcare systems. Notably, routine care still comprised 22 and 26% of campaigns primarily funding treatment in CAN and the UK, respectively, pointing to possible gaps despite universal healthcare. These gaps may be driven by rising out-of-pocket medication costs and insurance premiums in CAN and dissatisfaction with wait times for public care in the UK.^7,10,34^ Fundraising for experimental and approved but inaccessible therapies was the purpose of approximately 50% of campaigns in CAN and the UK highlighting a perceived shortcoming of publicly funded healthcare – insufficient or delayed access to novel and experimental treatments.

For cancer patients, financial hardship is common as novel expensive therapies become the standard of care and survival rates rise.^35–37^ Of the 9.5 million people diagnosed with cancer between 2000 and 2012 in the US, 42.4% had exhausted their life’s financial assets within two years^38^. The enormous scope of unmet financial need in cancer care is reflected in the marked overrepresentation of cancer campaigns on crowdfunding platforms.^4,7,14^ In our cohort, cancer accounted for 50-60% in all three countries. Notably, amongst cancer campaigns primarily funding treatment, campaigns for alternative therapies (22%) were at least twice as frequent as other diagnoses examined. Patients may seek alternative therapies for cancer to complement or replace proven treatments either by choice or because they were not available or failed.^13^ The popularity of alternative therapies for cancer creates the potential for unproven and potentially dangerous therapies yielding wasted resources, false hopes, delay of appropriate palliative care, and reduced survival.^13,16–18^

### Determinants of crowdfunding success

Beneficiary demographics, treatment type, the campaigning country, number of donors, fundraising goal, and campaign narrative length were important determinants of funding success. Initial regression analysis did not demonstrate a fundraising association with Facebook shares, but further sensitivity analysis that excluded donor numbers from the model, showed a strong association between Facebook shares and funds raised, reflecting moderate collinearity between Facebook shares and donor numbers. Previous crowdfunding research has shown that demographics^7,12^, donor numbers^39^, fundraising goal^11,12^, campaign narrative length^11,12,40^, and social media presence^4,7,40^ are predictors of crowdfunding success. Setting a higher goal for medical campaigns may indirectly communicate depth of need to donors and promote the concept of “deservingness.” Campaign narrative length reflects the importance of the illness narrative to funding success. Positive language in the narrative confers a fundraising advantage^12,40^ while references to the systemic injustices that led to a given MCF campaign are infrequently observed.^41^

### Limitations

This study has several limitations. First, while our dataset is larger than those in previous MCF research, it is only a small subset of popular MCF campaigns. Only campaigns visible on the “Discover” page of GoFundMe were included in our sample, which might have led to a bias towards more successful campaigns. We adopted his approach because it allowed us to filter out dummy or unverifiable campaigns that would have made our analysis unreliable. Further work is needed to explore trends in all campaigns and other factors affecting funding success.

Generalizability of our findings is limited by the use of only one MCF platform. It is difficult to truly ascertain the representation of MCF platforms due to a paucity of historical data and inaccessibility of proprietary information in a commercial market. Second, the veracity of the online data cannot be ensured, which restricts interpretability, but also highlights the important problems of misinformation, pseudoscience, and fraud in MCF. The lack of regulation and oversight raises questions about legal and medical responsibility. Third, there are inherent limitations in manual review, especially for demographic data using only media and textual context. However, in the absence of self-reported information, we believe this method provides an adequate granular view of the campaign information and demographics in MCF evidenced by near-perfect concordance between reviewers. Moreover, we believe our approach parallels the online crowdfunding experience of a potential donor who would rely on his/her perception of a beneficiary’s identity and attribute merit based on the illness narrative and media alone.

## Conclusions

We provide a foundational quantitative descriptive analysis of MCF and determinants of success in CAN, the UK, and the US. Despite MCF’s exponential growth, a vast majority of campaigns never meet their goal. We highlight important differences in MCF trends in publicly and privately funded healthcare systems, which point to unique gaps in healthcare funding and access in each setting. We also demonstrate racial and gender disparities in the use and success of MCF. MCF directly (through platforms that promote the victim narrative) and indirectly (by rewarding these narratives with funding success) promotes the myth that gaps in healthcare funding are due to misfortune and exceptionality rather than systemic failures. As such, MCF likely entrenches the systemic failures that potentiated its need. Thus, while crowdfunding has the potential to provide short-term relief from medical financial burden for a privileged subset of patients, it carries wider-reach paradoxical societal effect. Further research is needed to understand the social, ethical, and economic implications of MCF within each healthcare setting.

## Data Availability

The data that support the findings of this study are available upon request.

## Availability of data and materials

The data that support the findings of this study are available upon request.

## Authors’ contributions

Study concept and design: SS, EA, RM; Data acquisition and extraction: SS; Data manual review: EA, SS. Manual review adjudication: RM. Data analysis: SS; Interpretation of data: all authors; Manuscript preparation: SS, EA, CL, RM. All authors read and approved the final manuscript.

